# Does Physician Over-Service Improve the Quality of Care? A Standardised Patient Audit Study

**DOI:** 10.1101/2023.10.30.23297802

**Authors:** Yafei Si, Hazel Bateman, Shu Chen, Katja Hanewald, Bingqin Li, Min Su, Zhongliang Zhou

**Affiliations:** ARC Centre of Excellence in Population Ageing Research (CEPAR), The University of New South Wales, Sydney, Australia; School of Risk & Actuarial Studies, The University of New South Wales, Sydney, Australia; Social Policy Research Centre, The University of New South Wales, Sydney, Australia; School of Public Administration, Inner Mongolia University, Hohhot, China; School of Public Policy and Administration, Xi’an Jiaotong University, Xi’an, China

**Keywords:** quality of care, know-do gaps, physician over-service, standardised patient, primary care

## Abstract

Physicians’ “know-do gaps” are a key factor driving the poor quality of healthcare in many developing countries, but there is little guidance on how to address these gaps. We designed a standardised patient audit study in China to evaluate the impact of physician over-service on their investment in learning and disease management decisions. We find that physicians’ total over-service leads to a 19.2%, 15.6% and 10.8% significant increase in consultation length, adherence to checklists, and patient-centred communication, respectively, but no significant improvement in giving a correct diagnosis, drug prescription or referral. The effects on physicians’ investment in learning are driven by over-service in drug prescription rather than over-service in medical tests. Moreover, over-service in drug prescription significantly leads to a 28.0 percentage-point increase in the prescription of a correct drug. Our findings imply that physician over-service reduces their “know-do gaps” and improves healthcare quality despite the related inefficient use of medical resources.

**JEL classification:** D82; H75; I10; I11; I18; J45

## 1. Introduction

The quality of healthcare is poor in many low- and middle-income countries. For example, studies in Shaanxi, China found that physicians often spent less than 5 minutes on average on a patient consultation, completed only 18–38% of items on a checklist of recommended questions and tests, and scored only 27% on measures of patient-centred communication (Sylvia et al., 2015; Su et al., 2021). As a result, only 26% of clinical diagnoses and 36% of drug prescriptions were correct (Sylvia et al., 2015). Other developing countries, such as India, Ghana, Kenya, Vietnam and South Africa, face similar problems (Kwan et al., 2019; Zawahir et al., 2021).

Growing evidence suggests significant gaps between what physicians know how to do and what they do in practice (Das and Hammer, 2007; Gage et al., 2018; Leonard and Masatu, 2010; Mohanan et al., 2015), as a key factor driving the poor quality of healthcare in many developing countries. One common explanation is that a shortage of medical staff leads to an excessive workload and thus, physicians have insufficient time to complete all checklist items or be patient-centred (UNICEF, 2020). However, the “know-do gaps” can also signal that health systems fail to adequately motivate physicians to do their best to understand a patient’s problem because they are underpaid. The difference in causes between workload and motivation corresponds to a broad distinction between resource-orientated and incentive-based interventions. Resource-orientated policies demand a significant increase in the number of medical staff to reduce the average workload; For motivation-based policies, an intuitive intervention is to reward physicians with a higher market price to incentivise higher effort and thus improve the overall quality of care.^1^

However, due two key empirical challenges, few studies have explored the causal relationship between higher market prices, physicians’ extra effort, and healthcare quality. First, because a physician’s knowledge and experience often endogenously determine how much effort is exerted by him/her to guarantee basic quality requirements for a medical consultation, it is rare to observe an exogenous change in a physician’s market price which is not related this knowledge or experience. Second, higher quality of care can also lead to higher market prices. For example, physicians may hope to improve the accuracy of diagnosis and treatment by using more diagnostic tests, but this also drives up his/her market prices.

We designed a standardised patient (SP) audit study in primary care in China to overcome both of these empirical challenges by investigating the impact of physician over-service on healthcare quality. Standardised patients are acting patients trained to present symptoms and answer physicians’ questions in a consistent manner. The method helps us address the fundamental problems of both inferring healthcare quality and precisely identifying physician over-service.^2^ First, physicians often seek higher financial gains through over-service (Si et al., 2023), which is not necessarily related to their knowledge or competence. Therefore, we use the presence of physician over-service to proxy for the physician’s price changes under the fee-for-service financing system in primary care in China. Second, physicians’ tendency to over-service is predetermined by the hospital’s incentive structure. The decision to over-service is initiated when patients start their opening statements. Therefore, the associated changes in physicians’ effort and performance can be precisely identified. Similar to Mullainathan and Obermeyer, 2021, our identification strategy relies on the walk-in nature of primary hospitals in China. SPs arrive at random times and interact with random physicians.

Our study yields three main findings. First, we find that physicians’ total over-service improves their investment in learning, with a 19.2%, 15.6% and 10.8% significant increase in consultation length, adherence to checklists, and patient-centred communication, respectively. However, we find no significant improvement in disease management, such as giving a correct diagnosis, drug prescription or referral. Second, the effects on physicians’ investment in learning were driven by over-service in drug prescription rather than over-service in medical tests. Over-service in drug prescription significantly leads to a 28.0 percentage-point increase in the prescription of a correct drug and no effects on giving a correct diagnosis or referral. Third, physicians’ improved performance in drug prescription is mainly explained by the prescription of more drugs but not by physicians’ extra investment in learning.

This study adds evidence to three streams of literature. First, this study contributes to a large literature on variations in healthcare services and the value of these variations. The high prevalence of physician over-service is well documented in high-income countries across a wide range of services and is also increasingly recognised in low-income countries, which can cause physio-psychological harm and unnecessary out-of-pocket costs to patients (Brownlee et al., 2017). Despite the abundance of analyses of physician over-service in medical and economic research (Brownlee et al., 2017; Currie et al., 2014; Finkelstein et al., 2016), its impact on healthcare quality is poorly understood, and most prior studies could only assess the number of health services (Fang et al., 2021; Fowler et al., 2008; Si et al., 2020). Our main contribution is to demonstrate that physician over-service can also contribute to physicians’ learning and, thus, better healthcare in developing countries sharing the similar institutional setup for healthcare services as China. Failure to understand the causal relationship between physician over-service and healthcare quality can result in inappropriate policy interventions.

Second, this study contributes to a better understanding of the “know-do gaps” of physician practice. In a setting of China’s Shaanxi province in 2016, there were only 2.25 physicians per 1,000 population, fewer than the 4.4 qualified staff per 1,000 population recommended by the World Health Organization (WHO, 2016). However, even without structural changes in the allocation of medical staff, physician over-service can significantly improve physicians’ investment in learning and, thus, better performance in drug prescription. Adding to the previous research in India (Das et al., 2016), we explain that physicians may not reach their production possibility frontier in practice without adequate incentives, especially when there is no prior relationship between an agent and a principal, and the agent needs to learn about their customers’ problems and make decisions on their behalf (Dulleck and Kerschbamer, 2006). We conduct additional analysis and find no impact of workload on healthcare quality, which is consistent with Kovacs and Lagarde (2022). Therefore, in terms of health service delivery in China and other countries with similar concerns, it is equally, if not more, important for decision-makers to use effective incentives to motivate physicians to do their best by providing the most accurate care in contrast to the traditional method of increasing the number of medical staff.

Third, this study is closely related to the literature that documents the dynamics of physician decision-making. Physician over-service can improve healthcare quality in the short term, but its impacts on patients’ health outcomes can be very mixed in the long term. Physician over-service does not improve the accuracy of physicians’ decisions and, therefore, seems unlikely to improve their diagnostic skills via a *learning-by-doing* process over time. In contrast, the over-service may contribute to a rigid practice style. In particular, physicians can cultivate habits of prescription (Stern and Trajtenberg, 1998), with their favourite drugs accounting for more than 66% of the total prescriptions (Berndt et al., 2015). This kind of practice style often worsens physicians’ diagnostic skills over time. In this case, patients face a higher chance of taking the correct drug and the risk of polypharmacy as well. One reason for the lack of research in the field is the difficulty of precisely separating ‘unnecessary’ services from ‘high-quality’ services (Anderson and Chalkidou, 2008). Our methodological contribution using the SP method makes such research feasible in the future.

This paper proceeds as follows. In Section 2, we introduce the institutional context, the SP method, a measure for physician over-service and the statistical analysis. Section 3 describes the data, main results, and robustness tests. In Section 4, we discuss the findings and conclude.

## 2. Methods

### Institutional Context

The health system in China commonly uses a fee-for-service approach for both public and private hospitals, where the payments for consultation, medical tests and medications are unbundled. Primary hospitals in China provide walk-in services for patients without an appointment. In general, patients pay a fixed consultation fee for each visit. A physician has discretion over consultation length, the questions asked, the medical tests performed, and the drugs prescribed. Fees for medical tests and drugs are paid separately at the end of each visit (See *Figure 1*).

**Figure 1.**
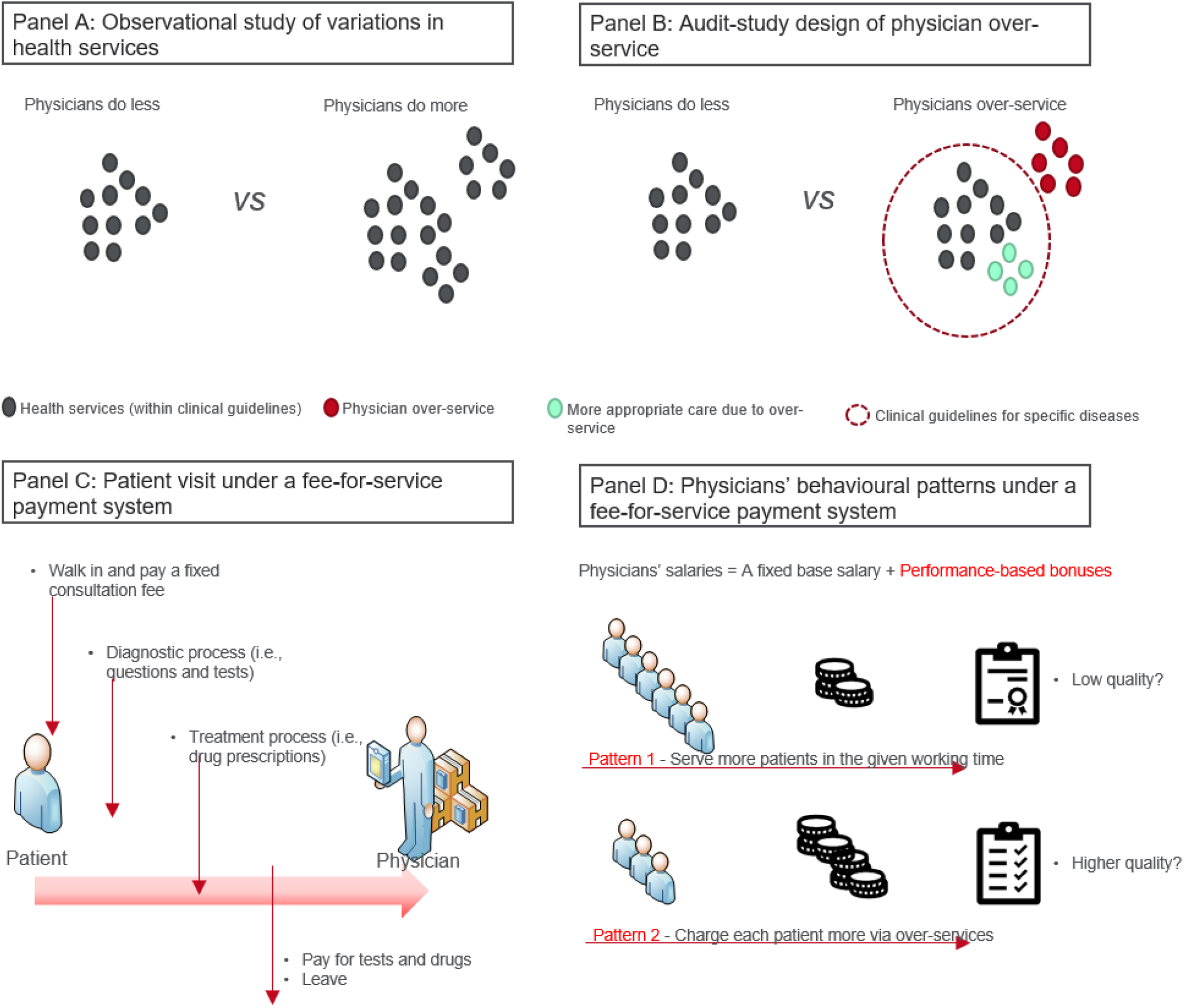
Theoretical and institutional context of the standardised patient audit study Note: *Panel A* illustrates an observational study design to investigate variations in health services. *Panel B* illustrates an audit study designed to investigate physician over-service. *Panel C* describes the institutional context of the standardised patient audit study. *Panel D* describes physicians’ behavioural patterns under a fee-for-service system.

Physicians’ salaries in China are determined by i) a fixed base salary based on qualifications and experience and ii) performance-based bonuses from the hospital. The performance-based bonus is generally determined by the number of patients served and the total earnings of the physician. However, usually, the bonus is much greater than base salary and departments rank physicians based on their performance so that the physicians are highly motivated to prescribe more drugs and diagnostic tests. The physicians’ payment structure under the fee-for-service system can result in a trade-off between the quantity and the quality of services provided. In theory physicians’ behaviour can have 2 patterns (See *Figure 1*).

For pattern 1, the incentive structure encourages physicians to see patients as quickly as possible, but this may compromise the quality of care. For pattern 2, physicians can decide to charge each patient more via over-service, which may not compromise or even improve the quality of care. An earlier study found that physician over-service led to a 118% increase in total cost compared to those without over-service (Si et al., 2023). The impact of over-service on the quality of care deserves prudent attention since the design of effective policies to regulate agents’ behaviour requires a good understanding of over-service per se and how it affects agents’ decision-making.

### Standardised Patients

We recruited 18 SPs from local communities and trained them to consistently present two chronic diseases: unstable angina and asthma. Three months before the audit study, physicians were informed about the SP visits but not the diseases to be tested. The SPs randomly visited physicians working in 63 primary hospitals in a capital city in western China on a workday in 2017 and 2018. We required SPs to choose the physician in the first office they passed after entering a hospital. Therefore, in theory, SPs and physicians were randomly paired because of the walk-in nature of primary care and the predetermined shift schedules of physicians.

SPs recorded comprehensive details about their interactions with the physicians using a structured questionnaire (i.e., consultation length, number of questions asked, medical tests ordered, diagnosis, medications, medical advice, and price charged). We instructed SPs to directly ask physicians for a diagnosis at the end of the visit if one had not yet been offered. SPs accepted and paid for all non-invasive medical tests and medications prescribed. Further details about the data collection process can be found in (Si et al., 2023). Our sample included 492 interactions between 169 physicians and 18 SPs.

The hospitals and physicians granted approval for the study three months before the visits. The study was approved by the Ethics Committee of the relevant universities.

### Quality of Healthcare

The first quality domain of interest is the physician’s investment in learning about a specific condition during a patient visit. We used three continuous variables.

- *Consultation length:* We used the length of direct physician-patient communication, excluding the time taken for tests, prescriptions, and dispensation of drugs (Das et al., 2016), measured in minutes. Since consultation is a crucial component of primary care practice, the length of a consultation is an important determinant of and proxy for provider effort in health care assessment (Bener et al., 2007; Elmore et al., 2016).
- *Adherence to checklists:* We measured the physicians’ adherence to an essential checklist of questions and tests for a specific disease in practice, which serves as the benchmark for correct diagnosis and drug prescription. This case-specific measure was defined as a count variable (maximum 20 for asthma and 22 for angina). The checklists for the two diseases presented by the SPs were based on two studies in India and China (Das et al., 2012; Sylvia et al., 2015).
- *Patient-centred communication:* We measured patient-centred communication in each physician-patient interaction using three components: exploring both the disease and the illness experience, understanding the whole person and finding common ground (Su et al., 2022). Patient-centred communication is known to be associated with reduced diagnostic tests and irrational patient requests without compromising patient satisfaction (Bertakis and Azari, 2011; Paterniti et al., 2010).

The second quality domain of interest is the effectiveness of disease management. We used three binary variables (0/1).

- *Correct diagnosis:* The SP method provides a standardised background and setting for the acting patients. Therefore, unambiguous clinical diagnoses should be provided after the visit if physicians follow the checklists. A diagnosis was classified as ‘correct’, ‘partially correct’ or ‘incorrect’ based on the guidelines on the checklist (Das et al., 2016; Sylvia et al., 2017). We created one binary variable to give the correct diagnosis for each interaction.
- *Correct drug prescription:* Drug prescriptions were considered ‘correct’ when there was at least one drug prescribed that would relieve/mitigate the underlying conditions. Other drug prescriptions were considered ‘incorrect’ (including unnecessary/harmful). A pilot study conducted with local physicians developed a list of ‘correct’ drugs for a prescription based on national treatment guidelines. We created one binary variable of giving the correct drug prescription for each interaction.
- *Referral:* This variable indicates whether the physician referred the SP to (or suggested that the SP visit) other (often higher-tier) hospitals after the visit. The interpretation of referral is not straightforward. Often, referrals are good, for example, when a physician refers a patient with a potentially life-threatening illness to a hospital. However, a referral may also reflect deflection of the case by the physician without treating the patient effectively. We included the referral variable to facilitate the interpretation of results and uncover the physicians’ decision-making process.

### Physician Over-service

The provision of unnecessary medical tests and drugs was identified based on the advice of a panel of medical experts (i.e., physicians, pharmacists and professors) and national clinical guidelines (Si et al., 2023). The panel classified each prescribed test and drug as essential/correct, unnecessary and even harmful; thus, we defined the presence of any unnecessary (or even harmful) tests/drugs as *physician over-service*. We created three binary variables for each interaction to measure over-service in medical tests, over-service in drugs, and the physician’s total over-service (in either tests or drugs).

### Statistical Analysis

For descriptive analysis, we explored the unadjusted association between quality metrics and physician over-service using the chi-squared test for binary variables and analysis of variance for continuous variables (Szklo and Nieto, 2014). Second, we used linear regression models to examine the relationship between physician over-service and the quality of care. The econometric specification was as follows:

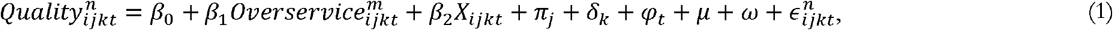

where 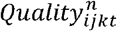 represents quality indicator *n* (i.e. consultation length, adherence to checklist, patient-centred communication, correct diagnosis, drug prescription and referral) for visit *i* at hospital *j* in district *k* on day *t*. 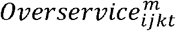 indicates the over-service in medical tests, the over-service in drug prescription or the total over-service. In the model, *X*_*ijkt*_ represents a set of observable physician characteristics and we further controlled for *π*_*j*_ hospital fixed effects, *μ* disease fixed effects and *ω* SP fixed effects. Robust standard errors were clustered at the hospital level.

We investigated two channels through which over-service is potentially linked to a higher quality of care. First, over-service *per se* can generate new information to help identify a disease (Currie and MacLeod, 2020); in addition, the provision of optimal care often entails experimentation and can involve some choices that turn out to have been sub-optimal *ex-post* (Currie and MacLeod, 2020). This process, known as *learning by doing*, can improve physicians’ diagnostic skills in the long term (Reese, 2011). Second, over-service can incentivise physicians to provide more appropriate care to assess and treat the disease (Das et al., 2016). The coefficients of physician over-service regarding the effectiveness of disease management include i) the effect of physician over-service *per se* and ii) the effect of providing more appropriate care within clinical guidelines. We separated the two using mediation analyses.

We performed a series of robustness tests. First, we excluded SP fixed effects in the regression models to confirm the comparability across SPs. Second, we constructed a propensity score to define high-propensity and low-propensity hospitals regarding physician over-service, using the *as-if* random variations in SPs’ arrival time to establish robust causal evidence. Third, we varied the thresholds defining high-propensity hospitals (i.e., 40%, 50% and 60%). Fourth, we used an instrumental variable (IV) strategy to evaluate the impact of physician over-service on healthcare quality. We report two-sided *p*-values (unadjusted for multiple comparisons) and 95% CI throughout this study. A *p*-value < 0.05 was considered to indicate statistical significance. All analyses were performed using Stata 16.0 (Stata Corp LLP, College Station, TX, USA).

## 3. Results

Overall, physicians spent 6.21 (95% confidence interval (CI): 5.80 to 6.61) minutes per patient for a consultation, completed 7.12 (95% CI: 6.83 to 7.41; out of 20/22) items in the checklist and scored 23.22 (95% CI: 22.66 to 23.77; out of 84) in patient-centred communication. In addition, 44.11% (95% CI: 39.70% to 48.51%) of SP visits had a correct diagnosis, 11.38% (95% CI: 8.57% to 14.20%) had a correct drug prescription, and 41.61% (95% CI: 33.30% to 49.91%) resulted in a referral. We found that 54.67% of the SP visits involved over-service in medical tests and 28.05% involved over-service in drugs. Overall, 72.15% of the SP visits involved over-service either in medical tests or in drugs.

### The Impact of Physician Over-service

We began with an exploratory analysis to investigate the relationship between physician over-service and quality metrics using a bivariate analysis (*Figure 2*). Overall, physicians’ investment in learning was higher in the presence of physician over-service. However, this greater investment in learning in the over-service group did not consistently translate into better performance in disease management. For example, the rate of giving a correct diagnosis was lower, but the rate of giving a correct drug prescription was significantly higher in the over-service group than in its counterpart group. We then described the quality metrics according to over-service in medical tests or drugs (*Figure S1*). The patterns we observed did not change substantially.

**Figure 2.**
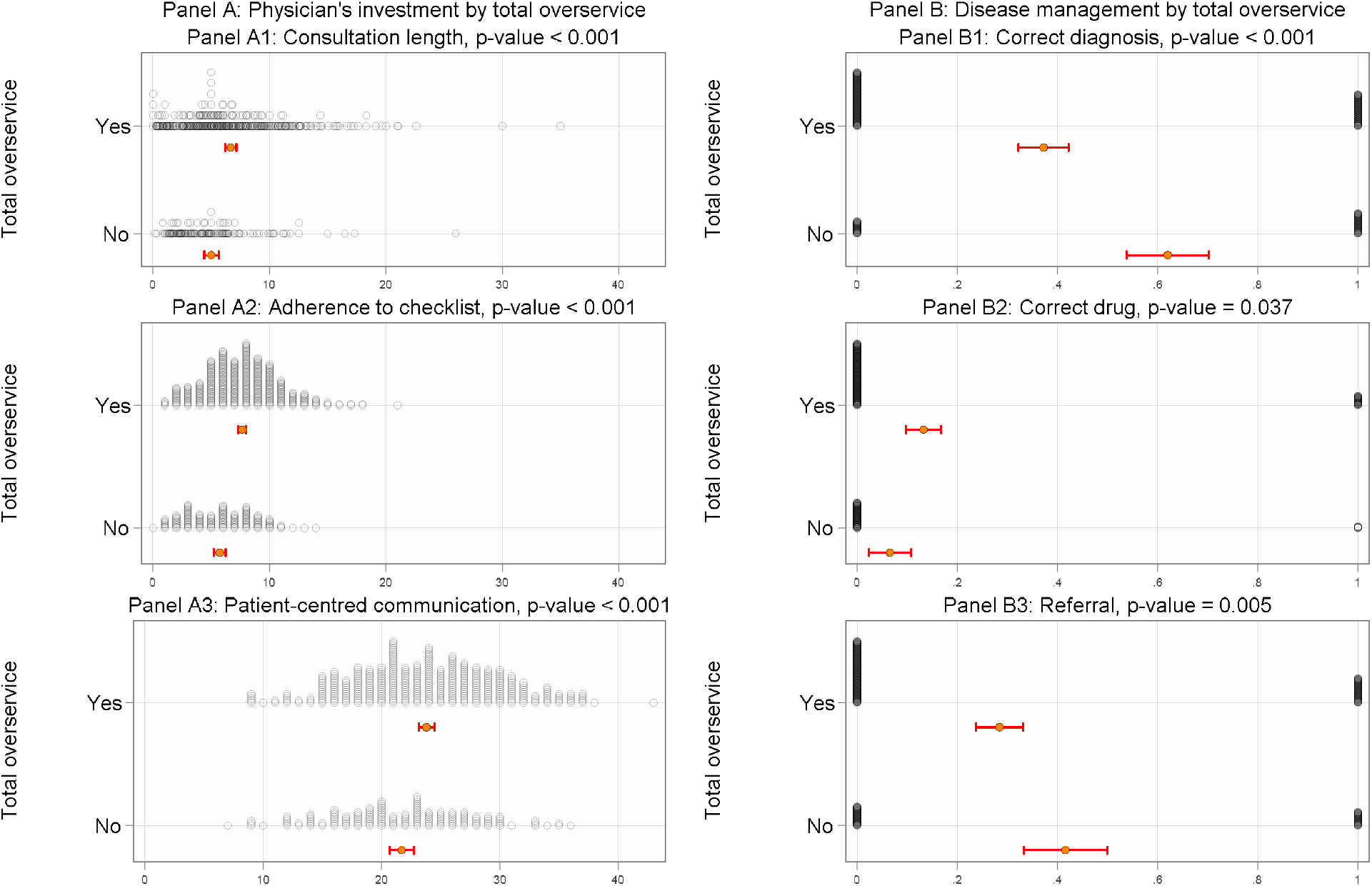
Relationship between physician over-service and quality of care Note: We tested for statistical differences using the chi-squared test for binary variables (consultation length) and analysis of variance for continuous variables (adherence to checklist and patient-centred communication). The figure reports the mean and 95% confidence intervals for all quality metrics and shows the distribution of all observations.

We used econometric models to examine the relationship between physician over-service and healthcare quality after controlling for physician characteristics and a series of fixed effects. Physicians’ total over-service was associated with significantly better learning (*Figure 3 Panel A*). For example, excluding the time when patients were sent for medical tests, physicians’ total over-service was significantly associated with a 0.96-minute increase (95% CI: 0.03 to 1.90) in consultation length, a 0.91-item increase (95% CI: 0.27 to 1.56) in adherence to checklists and a 2.38-unit increase (95% CI: 0.79 to 3.96) in patient-centred communication compared to those without over-service. This represented a 19.20%, 15.85% and 10.76% increase in consultation length, adherence to checklists and patient-centred communication, respectively. However, we found that physicians’ total over-service was not significantly associated with any change in giving a correct diagnosis or correct drug prescriptions. Physicians’ total over-service was weakly associated with an 11.5 percentage-point decrease (95% CI: 2.0 to 25.1) in referrals, with statistical significance at the 10% level.

**Figure 3.**
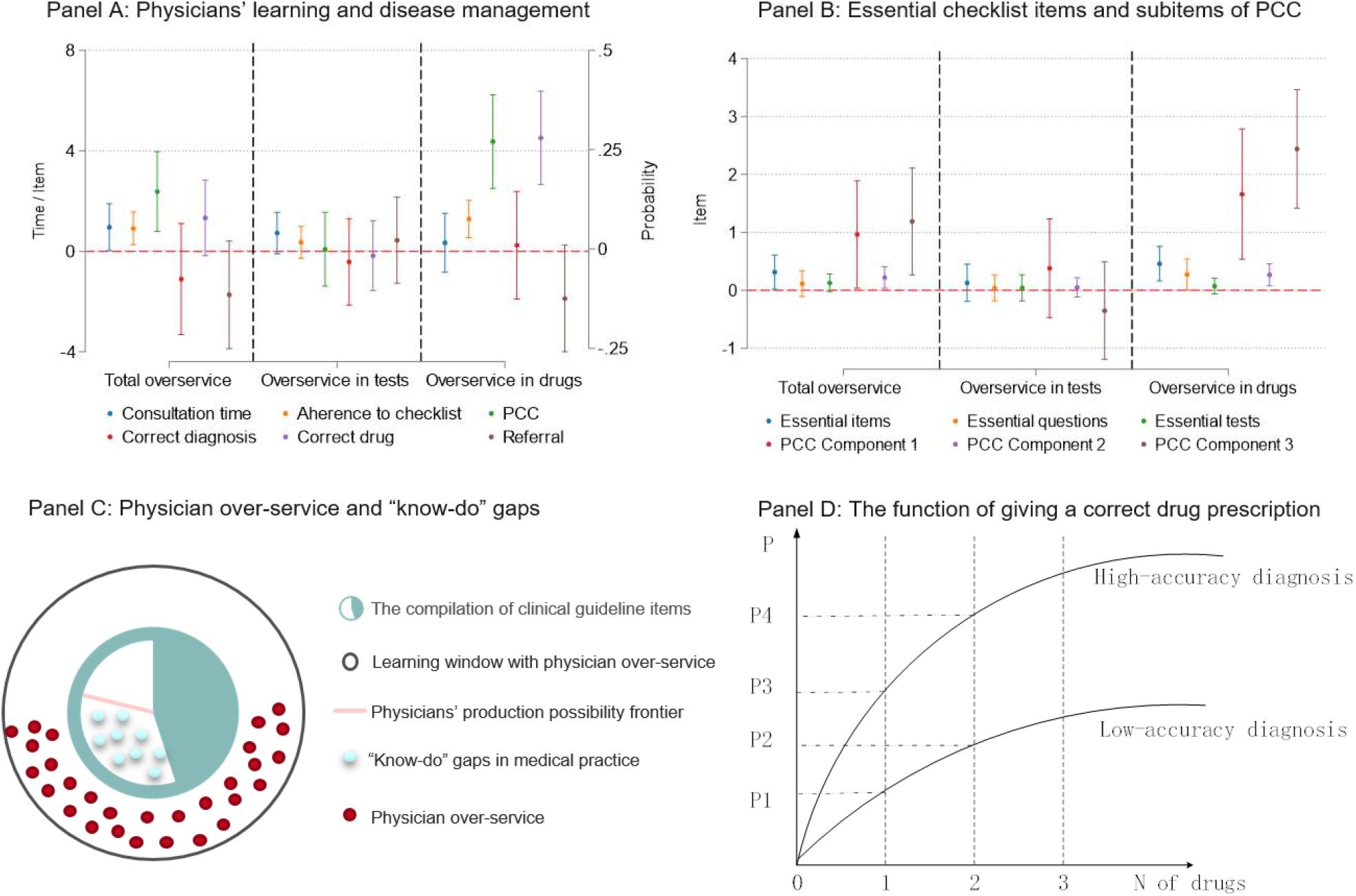
Association between physician over-service and the quality of care Note: *Panel A* shows the estimates of our econometric specification 1. Point estimates and their 95% confidence intervals are plotted. PCC denotes patient-centred communication. *Panel B* presents the estimates of our econometric model for essential items in checklists and sub-items of PCC. *Panel C* shows the conceptualisation of physicians’ production possibility frontier and ‘know-do’ gaps in medical practice. *Panel D* depicts how the accuracy of diagnosis and the number of drugs prescribed (N) jointly determine the cumulative probability of giving a correct drug prescription (P).

Interestingly, physician performance diverged across over-service in medical tests and drugs. First, over-service in medical tests was not significantly associated with any change in learning investment or in disease management (*Figure 3 Panel A*). However, over-service in drugs was associated with a significant increase in adherence to checklists (1.29 items, 95% CI: 0.55 to 2.03), the patient-centred communication score (4.37 units, 95% CI: 2.51 to 6.23) and giving a correct drug prescription (28.0 percentage points, 95% CI: 16.2 to 39.7). Although there was no improvement in consultation length or giving a correct diagnosis, we also found a 12.5 percentage-point decrease (95% CI: 0.9 to 25.9) in referrals.

To explain the confusion of better learning but unchanged disease management, we further examined the impact of physician over-service on essential items in the checklists (Das et al., 2016) and three sub-components of patient-centred communication (Su et al., 2021). The information provided by essential items is more important than that by common items for physicians to give a correct diagnosis and drug prescription. We found that physicians’ total over-service was statistically associated with a significant increase in adherence to essential items in the checklist and all three sub-components of patient-centred communication (*Figure 3 Panel C*). The effect size was bigger when we considered over-service in drugs but not in medical tests (*Table S3*).

To separate the information on physician over-service *per se* from that on more appropriate care due to physician over-service, we performed mediation analyses by adding the three metrics of physicians’ investment in learning to statistical models to explain the effectiveness of disease management. Our results remained consistent for physicians’ total over-service and over-service in medical tests. However, over-service in drugs was associated with a significant increase (23.6 percentage points, 95% CI: 11.3 to 35.8) in giving a correct drug prescription, meaning that the improved performance in drug prescription was mainly explained by over-service in drugs *per se*. Again, we found no significant change in the accuracy of diagnosis (*Table S4*).

### Robustness tests

We performed four theory-based robustness tests, none of which altered our results substantially. First, to examine whether our SPs were highly comparable to each other, we excluded patient fixed effects in the model (*Table S5*) and found good internal validity of the study. Second, to address potential endogeneity, since physician knowledge and motivation were not observed in the model, we constructed the probability of physician over-service in one hospital when an SP arrived, and whether or not an SP was assigned to visit a hospital where physicians had a high propensity for over-service was *as-if* random. On average, each hospital received 7.81 SP visits. Different SPs were randomly sent to visit the same hospital. The propensity for over-service followed approximately normal distributions (*Figure S2*). We compared quality metrics in two types of hospitals, that is, those with a high and low propensity for physician over-service, with high-propensity hospitals (50% and higher) as the treatment group and low-propensity hospitals (lower than 50%) as the control group. Our results were robust (*Table S6*), although the statistical estimation was less efficient because we could not control for hospital fixed effects. Third, our results did not change much when we used a variety of thresholds (i.e., 40% and 60%) to define high-propensity hospitals (*Table S7*). Fourth, our results remained very consistent when using an instrumental variable strategy to estimate the effect of physician over-service (*Table S8*).

### Workload and Healthcare Quality

Consistent with Kovacs and Lagarde (2022), we used the number of patients waiting to see the same provider at the hospital when SPs arrived as a simple measure of workload or patient load. Since each hospital received 7.81 SP visits on average, we also captured the average levels of workload in a hospital to describe how busy a hospital was when an SP arrived. We created a dummy variable equal to one (busy time) when the number of patients waiting to see the same provider at the hospital when the SP arrived exceeds the number of patients at its “average level”, or equal to zero (quiet time). Whether or not an SP was assigned to visit a hospital at its busy time was *as-if* random. We find no significant changes in consultation length, adherence to checklists, patient-centred communication, giving a correct diagnosis, correct drug prescription or referral (*Tabe S9*). Our results suggest that the workload does not necessarily relate to the healthcare quality.

## 4. Discussion and Conclusion

We present audit-study evidence on the impact of physician over-service on the provision of some inexpensive but potentially lifesaving diagnoses and treatments in a primary care setting in China. Our findings suggest that physician over-service improves health care quality under a fee-for-service financing system in primary care in China. Our findings support the conclusion that physicians are less motivated and do not reach their production possibility frontier in practice in developing countries (Kovacs and Lagarde, 2022). Physician over-service can, at least partly, attenuate their ‘know-do’ gaps (see *Figure 3 Panel C*).

It is surprising that physicians’ better learning associated with over-service did not improve the accuracy of their decisions on disease management, although we found a higher chance of correct drug prescriptions associated with physician over-service. Physicians’ performance in drug prescriptions is, in theory, a function of 1) the number of drugs prescribed and 2) the average probability of a prescribed drug being correct (details in *Supplement 1*). Therefore, physicians can perform better in drug prescriptions based on a higher-accuracy diagnosis, an indicator with no improvement in the study. In contrast, the simple strategy of prescribing more drugs can significantly improve physicians’ performance in the cumulative probability of giving a correct drug prescription even though the accuracy of diagnosis is very low (*Figure 3 Panel D*).

We further investigated why physician over-service did not improve the accuracy of their decisions on disease management. Basically, physicians’ better learning would not matter if the increase is marginal, only associated with unimportant information, or not patient-centred enough. However, the magnitude of the increase in learning was reasonably moderate or strong, physicians considered important information by addressing several essential items on the checklist, and physicians’ better learning was captured in all sub-components of patient-centred communication. The abovementioned explanations are not exhaustive, but it is reasonable to argue that in the study, physicians seem less responsive to new and important information gained from practice. This is clearly explained using an iterative *Bayesian learning* model (*Figure 4*).

**Figure 4.**
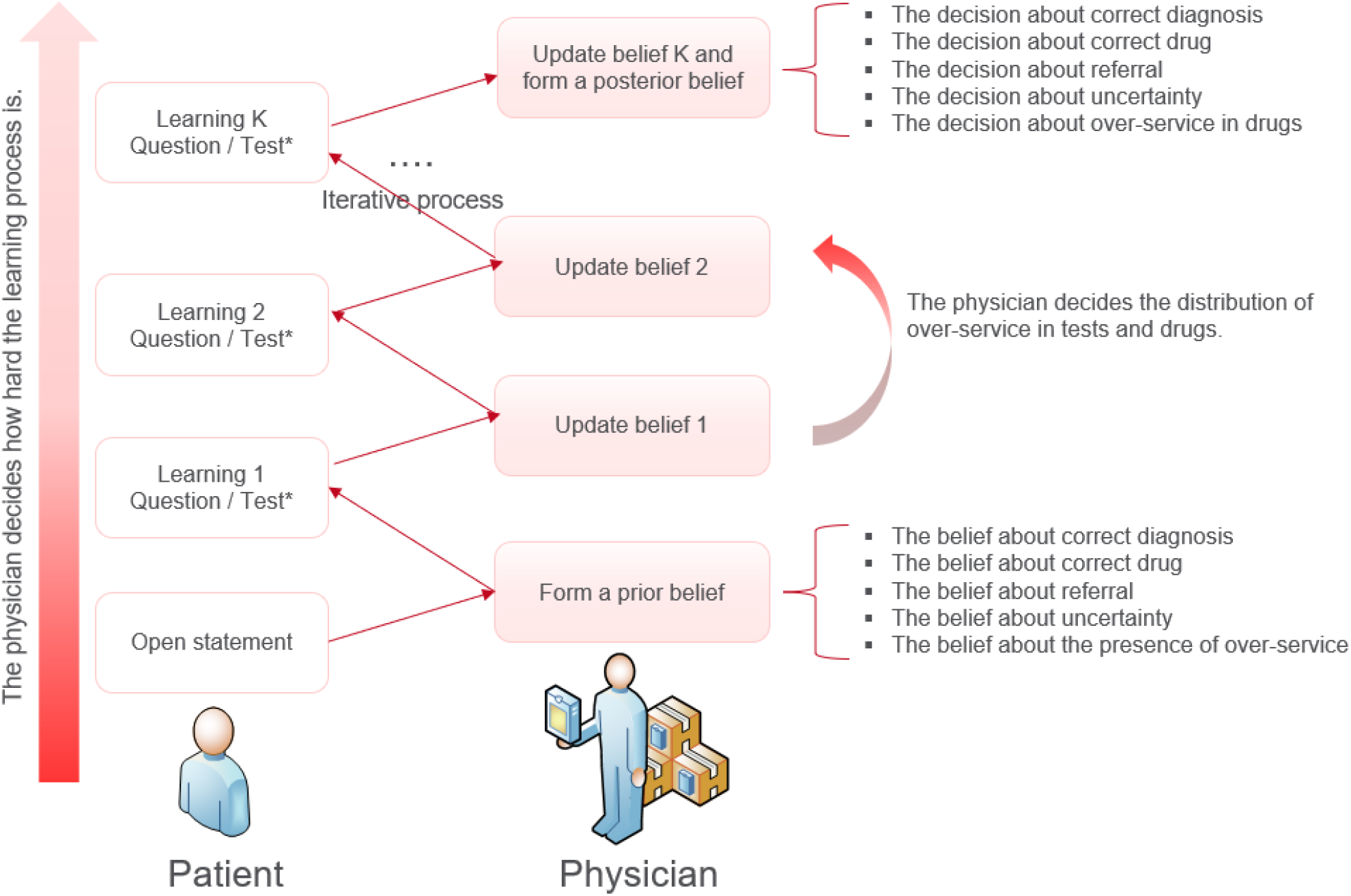
Iterative Bayesian learning process Note: The model is derived from Das et al. (2016). Test* denotes either a recommended or an unnecessary medical test. In the beginning, the physician forms a prior belief about the true illness and decides whether to over-provide services after the patient’s opening statement. The physician will not be penalized for poor quality of care since his/her salary is not linked to performance. The physician will have direct financial rewards related to the over-service. The physician can fully choose how many interactions to conduct (k = 1…. K) and form his/her posterior belief about the true illness of the patient. The diagnosis could be more precise with more iterations. Since over-services in medical tests and drugs are partial substitutes (Si et al. 2023), the physician balances the distribution of these over-service items, but this does not necessarily affect his/her discretion regarding the patient’s true illness and the correct drug prescription.

The effects of over-service in drugs but not in medical tests on physicians’ better learning can be explained in three ways. First, adherence to checklists and over-service in tests are partial substitutes, which can offset the effect of over-service in medical tests on physicians’ investment in learning compared with the effect of over-service in drugs. Second, the physician’s learning process for a specific patient can be temporarily interrupted when patients are sent to undergo medical tests because the physician usually initiates another consultation with the next patient in the queue (Wu, 2019; Zhu et al., 2021). Third, because SPs did not pay for all invasive tests.

Our results were robust to several robustness tests, and the effect identified in the study can be interpreted as causal when we use the propensity score for over-service to confirm the validity of our results. However, we acknowledge two limitations. First, we only discussed physician decision-making in primary care using two common conditions, and our results may not be generalisable to critical illness or tertiary care. Primary care is usually an essential first step in identifying the need for tertiary care and directing patients accordingly, and the benefits of physician over-service could outweigh its harms in the short term. Therefore, physicians were less likely to refer patients to other providers in the presence of over-service. Second, we used objective quality metrics, which are not necessarily equal to patients’ perceptions. How patients’ perceptions converge and diverge with physicians’ objective behaviours is beyond the scope of this study. Patients may prefer to choose physicians with specific characteristics (Das et al., 2016), and the preference, in turn, is likely to account for the physicians’ decision-making process.

## Data availability

The study was approved by the Ethics Committee of Xi’an Jiaotong University Health Science Centre (Approval number: 2015-406) and the Ethics Committee of the University of New South Wales (HC210354). The data are not publicly available due to restrictions of the ethics approval for this study. The code scripts used in this analysis are available from the corresponding authors upon reasonable request.

## Acknowledgements

The authors are grateful to all standardised patients and student instructors for collecting the data. YS framed the research question, contributed to data management and analysis and drafted the first version of the paper; SC and MS helped frame the research question and analytical approach and contributed to the drafting of the paper; MS verified the data. KH, BL, HB and ZZ oversaw the project, contributed to the framing of the research question and drafted significant portions of the paper. The authors appreciate very helpful comments from Prof Michael Keane, Prof Denzil Fiebig, Prof Anthony Scott, Prof Xi Chen, Prof Gang Chen, Dr Guy Mayraz, Dr Sophie Yan, and participants of Risk and Actuarial Studies seminars at UNSW Sydney, AHED 2021, and AHES 2023. All authors approved the final version of the paper.

## Footnotes

1 For the market price of a physician, we mean physicians’ total financial gains through market activities. In general, the total financial gains include basic salary and performance-based bonus in many low-and-middle income countries.

2 Over-service is defined as ‘to serve someone or something unnecessarily or to an excessive degree’.

