## Supplementary information for "Does Physician Over-Service Improve the Quality of Care? A Standardised Patient Audit Study"

[Table S6 Comparing the quality of care across high-propensity (>=50%) and low-propensity (<50%) hospitals	9](#_Toc145523482)

### Figure S1 Distribution of quality of care by over-service in medical tests and drugs

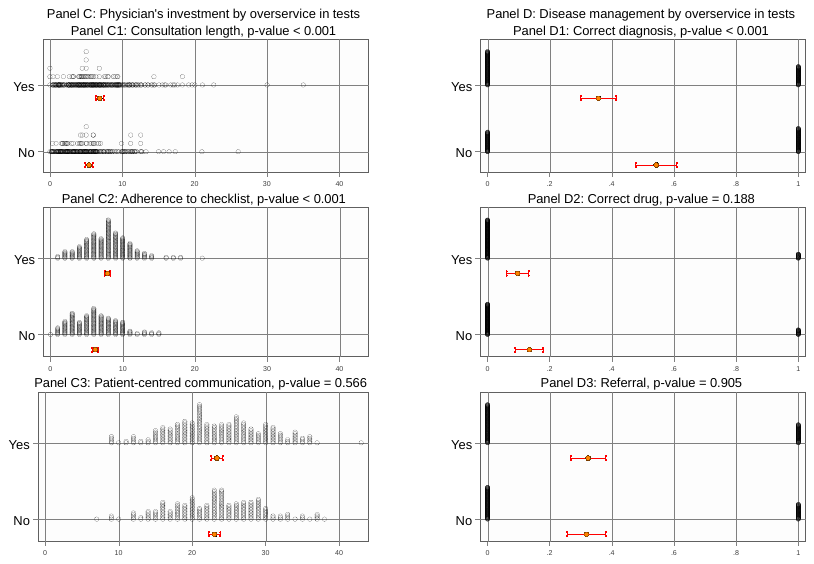

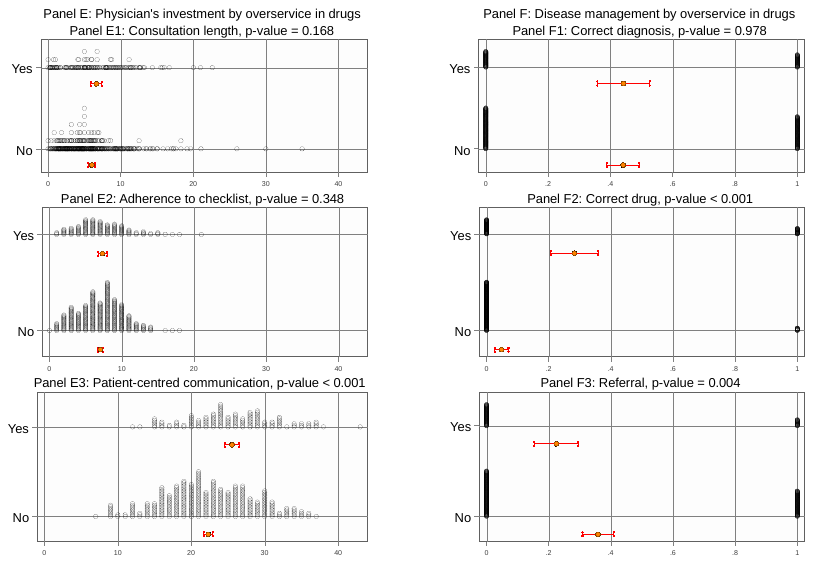

### Table S1 Summary statistics

|  | (1) |  |  | (2) |  |  | (3) |  |  |  |
| --- | --- | --- | --- | --- | --- | --- | --- | --- | --- | --- |
|  | Total over-service | |  | Over-service in tests | |  | Over-service in drugs | |  |  |
|  | No | Yes | p-value | No | Yes | p-value | No | Yes | p-value | Overall |
| Consultation time | 5.01 | 6.67 | <0.001 | 5.40 | 6.88 | <0.001 | 6.03 | 6.66 | 0.168 | 6.21 |
|  | [4.38, 5.63] | [6.18, 7.16] |  | [4.88, 5.91] | [6.29, 7.46] |  | [5.56, 6.50] | [5.90, 7.42] |  | [5.80, 6.61] |
| Adherence to checklist | 5.74 | 7.65 | <0.001 | 6.20 | 7.88 | <0.001 | 7.03 | 7.34 | 0.348 | 7.12 |
|  | [5.24, 6.23] | [7.31, 7.99] |  | [5.78, 6.61] | [7.50, 8.26] |  | [6.70, 7.36] | [6.73, 7.95] |  | [6.83, 7.41] |
| PCC | 21.69 | 23.81 | <0.001 | 23.04 | 23.36 | 0.566 | 22.31 | 25.54 | <0.001 | 23.22 |
|  | [20.68, 22.70] | [23.16, 24.46] |  | [22.24, 23.84] | [22.60, 24.13] |  | [21.68, 22.95] | [24.53, 26.54] |  | [22.66, 23.77] |
| Correct diagnosis | 0.6204 | 0.3718 | <0.001 | 0.5426 | 0.3569 | <0.001 | 0.4407 | 0.4420 | 0.978 | 0.4411 |
|  | [0.5387, 0.7022] | [0.3214, 0.4223] |  | [0.4769, 0.6083] | [0.2994, 0.4144] |  | [0.3888, 0.4926] | [0.3587, 0.5254] |  | [0.3970, 0.4851] |
| Correct drug | 0.0657 | 0.1324 | 0.037 | 0.1345 | 0.0967 | 0.188 | 0.0480 | 0.2826 | <0.001 | 0.1138 |
|  | [0.0240, 0.1074] | [0.0970, 0.1678] |  | [0.0895, 0.1795] | [0.0612, 0.1321] |  | [0.0257, 0.0704] | [0.2070, 0.3582] |  | [0.0857, 0.1420] |
| Referral | 0.4161 | 0.2845 | 0.005 | 0.3184 | 0.3234 | 0.905 | 0.3588 | 0.2246 | 0.004 | 0.4161 |
|  | [0.3330, 0.4991] | [0.2374, 0.3316] |  | [0.2570, 0.3798] | [0.2673, 0.3796] |  | [0.3086, 0.4089] | [0.1546, 0.2947] |  | [0.3330, 0.4991] |

Note: p-values were calculated using analysis of variance (ANOVA) for continuous variables and chi-squared tests for binary variables; PCC denotes patient-centred communication. ^*^ *p* < .1, ^**^ *p* < .05, ^***^ *p* < .01

### Table S2 Association between physician over-service and quality of care

|  | (1) | (2) | (3) | (4) | (5) | (6) | (7) | (8) | (9) |
| --- | --- | --- | --- | --- | --- | --- | --- | --- | --- |
|  | Time | Checklist | PCC | Time | Checklist | PCC | Time | Checklist | PCC |
| Total over-service | 0.961^**^ | 0.914^***^ | 2.376^***^ |  |  |  |  |  |  |
|  | [0.025, 1.897] | [0.269, 1.560] | [0.789, 3.963] |  |  |  |  |  |  |
| Over-service in tests |  |  |  | 0.727^*^ | 0.361 | 0.082 |  |  |  |
|  |  |  |  | [-0.099,1.553] | [-0.273,0.994] | [-1.386,1.550] |  |  |  |
| Over-service in drugs |  |  |  |  |  |  | 0.338 | 1.286^***^ | 4.369^***^ |
|  |  |  |  |  |  |  | [-0.835,1.512] | [0.547,2.026] | [2.509,6.230] |
| *N* | 492 | 492 | 492 | 492 | 492 | 492 | 492 | 492 | 492 |
| adj. *R*^2^ | 0.23 | 0.47 | 0.22 | 0.23 | 0.46 | 0.20 | 0.22 | 0.48 | 0.27 |

|  | (1) | (2) | (3) | (4) | (5) | (6) | (7) | (8) | (9) |
| --- | --- | --- | --- | --- | --- | --- | --- | --- | --- |
|  | Diagnosis | Treatment | Referral | Diagnosis | Treatment | Referral | Diagnosis | Treatment | Referral |
| Total over-service | -0.076 | 0.078 | -0.115^*^ |  |  |  |  |  |  |
|  | [-0.215, 0.064] | [-0.017, 0.174] | [-0.251, 0.020] |  |  |  |  |  |  |
| Over-service in tests |  |  |  | -0.033 | -0.017 | 0.022 |  |  |  |
|  |  |  |  | [-0.142, 0.076] | [-0.105, 0.071] | [-0.086, 0.130] |  |  |  |
| Over-service in drugs |  |  |  |  |  |  | 0.009 | 0.280^***^ | -0.125^*^ |
|  |  |  |  |  |  |  | [-0.126, 0.145] | [0.162, 0.397] | [-0.259, 0.009] |
| *N* | 492 | 492 | 492 | 492 | 492 | 492 | 492 | 492 | 492 |
| adj. *R*^2^ | 0.19 | -0.00 | 0.18 | 0.19 | -0.01 | 0.17 | 0.19 | 0.10 | 0.18 |

Note: Covariates included physician gender and age. County, hospital, day of week, month, year, case, and patient fixed effect were controlled for in the regression. Robust standard errors were clustered at the hospital level. PCC denotes patient-centred communication. 95% confidence intervals in brackets. ^*^ *p* < .1, ^**^ *p* < .05, ^***^ *p* < .01

### Table S3 Subitems of checklist and PCC

|  | (1) | (2) | (3) | (4) | (5) | (6) | (7) | (8) | (9) |
| --- | --- | --- | --- | --- | --- | --- | --- | --- | --- |
| Checklist | Item | Question | Examination | Item | Question | Examination | Item | Question | Examination |
| Total over-service | 0.315^**^ | 0.114 | 0.129^*^ |  |  |  |  |  |  |
|  | [0.021, 0.609] | [-0.108, 0.337] | [-0.022, 0.279] |  |  |  |  |  |  |
| Over-service in tests |  |  |  | 0.130 | 0.042 | 0.042 |  |  |  |
|  |  |  |  | [-0.191, 0.452] | [-0.182, 0.266] | [-0.182, 0.266] |  |  |  |
| Over-service in drugs |  |  |  |  |  |  | 0.459^***^ | 0.274^**^ | 0.072 |
|  |  |  |  |  |  |  | [0.163, 0.756] | [0.009, 0.539] | [-0.064, 0.208] |
| *N* | 492 | 492 | 492 | 492 | 492 | 492 | 492 | 492 | 492 |
| adj. *R*^2^ | 0.16 | 0.13 | 0.11 | 0.15 | 0.13 | 0.13 | 0.17 | 0.14 | 0.10 |

|  | (1) | (2) | (3) | (4) | (5) | (6) | (7) | (8) | (9) |
| --- | --- | --- | --- | --- | --- | --- | --- | --- | --- |
| PCC | C1 | C2 | C3 | C1 | C2 | C3 | C1 | C2 | C3 |
| Total over-service | 0.965^**^ | 0.222^**^ | 1.189^**^ |  |  |  |  |  |  |
|  | [0.039, 1.891] | [0.039, 0.405] | [0.269, 2.110] |  |  |  |  |  |  |
| Over-service in tests |  |  |  | 0.382 | 0.051 | -0.351 |  |  |  |
|  |  |  |  | [-0.471, 1.236] | [-0.116, 0.218] | [-1.192, 0.489] |  |  |  |
| Over-service in drugs |  |  |  |  |  |  | 1.660^***^ | 0.268^***^ | 2.441^***^ |
|  |  |  |  |  |  |  | [0.537, 2.783] | [0.079, 0.457] | [1.418, 3.465] |
| *N* | 492 | 492 | 492 | 492 | 492 | 492 | 492 | 492 | 492 |
| adj. *R*^2^ | 0.32 | 0.17 | 0.24 | 0.32 | 0.15 | 0.22 | 0.34 | 0.18 | 0.29 |

Note: Covariates included physician gender and age. County, hospital, day of week, month, year, case, and patient fixed effect were controlled for in the regression. Robust standard errors were clustered at the hospital level. PCC denotes patient-centred communication. 95% confidence intervals in brackets. ^*^ *p* < .1, ^**^ *p* < .05, ^***^ *p* < .01

### Table S4 Mediation analysis

|  | (1) | (2) | (3) | (4) | (5) | (6) | (7) | (8) | (9) |
| --- | --- | --- | --- | --- | --- | --- | --- | --- | --- |
|  | Diagnosis | Treatment | Referral | Diagnosis | Treatment | Referral | Diagnosis | Treatment | Referral |
| Total over-service | -0.110 | 0.046 | -0.121^*^ |  |  |  |  |  |  |
|  | [-0.244, 0.024] | [-0.046, 0.139] | [-0.257, 0.016] |  |  |  |  |  |  |
| Over-service in tests |  |  |  | -0.012 | -0.015 | 0.032 |  |  |  |
|  |  |  |  | [-0.113, 0.089] | [-0.095, 0.065] | [-0.076, 0.140] |  |  |  |
| Over-service in drugs |  |  |  |  |  |  | -0.085 | 0.236^***^ | -0.156^**^ |
|  |  |  |  |  |  |  | [-0.222, 0.052] | [0.113, 0.358] | [-0.292, -0.021] |
| Time | -0.007 | -0.000 | -0.005 | -0.008 | 0.000 | -0.006 | -0.008 | 0.000 | -0.006 |
|  | [-0.018, 0.004] | [-0.007, 0.007] | [-0.016, 0.005] | [-0.019, 0.004] | [-0.007, 0.007] | [-0.017, 0.004] | [-0.020, 0.004] | [-0.006, 0.007] | [-0.017, 0.004] |
| Checklist | -0.049^***^ | -0.011 | -0.015 | -0.050^***^ | -0.010 | -0.017 | -0.050^***^ | -0.009 | -0.017 |
|  | [-0.077, -0.021] | [-0.030, 0.008] | [-0.042, 0.012] | [-0.077, -0.022] | [-0.029, 0.008] | [-0.043, 0.009] | [-0.078, -0.022] | [-0.027, 0.008] | [-0.043, 0.009] |
| PCC | 0.036^***^ | 0.018^***^ | 0.010^*^ | 0.035^***^ | 0.018^***^ | 0.009 | 0.037^***^ | 0.013^***^ | 0.013^**^ |
|  | [0.025, 0.047] | [0.010, 0.026] | [-0.001, 0.022] | [0.024, 0.046] | [0.010, 0.026] | [-0.002, 0.021] | [0.025, 0.049] | [0.005, 0.021] | [0.001, 0.024] |
| *N* | 492 | 492 | 492 | 492 | 492 | 492 | 492 | 492 | 492 |
| adj. *R*^2^ | 0.26 | 0.06 | 0.18 | 0.25 | 0.05 | 0.17 | 0.26 | 0.13 | 0.18 |

Note: Covariates included physician gender and age. County, hospital, day of week, month, year, case, and patient fixed effect were controlled for in the regression. Robust standard errors were clustered at the hospital level. PCC denotes patient-centred communication. 95% confidence intervals in brackets. ^*^ *p* < .1, ^**^ *p* < .05, ^***^ *p* < .01

### Table S5 Excluding patient fixed effect

|  | (1) | (2) | (3) | (4) | (5) | (6) | (7) | (8) | (9) |
| --- | --- | --- | --- | --- | --- | --- | --- | --- | --- |
|  | Time | Checklist | PCC | Time | Checklist | PCC | Time | Checklist | PCC |
| Total over-service | 1.031^**^ | 1.030^***^ | 2.493^***^ |  |  |  |  |  |  |
|  | [0.175, 1.888] | [0.427, 1.633] | [0.915, 4.072] |  |  |  |  |  |  |
| Over-service in tests |  |  |  | 1.097^***^ | 0.575^*^ | 0.263 |  |  |  |
|  |  |  |  | [0.296, 1.898] | [-0.099, 1.249] | [-1.272, 1.799] |  |  |  |
| Over-service in drugs |  |  |  |  |  |  | 0.172 | 1.128^***^ | 4.029^***^ |
|  |  |  |  |  |  |  | [-1.010, 1.355] | [0.381, 1.875] | [2.273, 5.785] |
| *N* | 492 | 492 | 492 | 492 | 492 | 492 | 492 | 492 | 492 |
| adj. *R*^2^ | 0.12 | 0.41 | 0.17 | 0.12 | 0.40 | 0.14 | 0.11 | 0.41 | 0.21 |

|  | (1) | (2) | (3) | (4) | (5) | (6) | (7) | (8) | (9) |
| --- | --- | --- | --- | --- | --- | --- | --- | --- | --- |
|  | Diagnosis | Treatment | Referral | Diagnosis | Treatment | Referral | Diagnosis | Treatment | Referral |
| Total over-service | -0.073 | 0.072 | -0.116^*^ |  |  |  |  |  |  |
|  | [-0.210, 0.064] | [-0.018, 0.163] | [-0.252, 0.020] |  |  |  |  |  |  |
| Over-service in tests |  |  |  | -0.047 | -0.030 | 0.021 |  |  |  |
|  |  |  |  | [-0.154, 0.060] | [-0.116, 0.056] | [-0.089, 0.130] |  |  |  |
| Over-service in drugs |  |  |  |  |  |  | 0.012 | 0.230^***^ | -0.077 |
|  |  |  |  |  |  |  | [-0.102, 0.126] | [0.132, 0.329] | [-0.197, 0.042] |
| *N* | 492 | 492 | 492 | 492 | 492 | 492 | 492 | 492 | 492 |
| adj. *R*^2^ | 0.19 | 0.00 | 0.18 | 0.19 | -0.01 | 0.17 | 0.03 | 0.10 | 0.13 |

Note: Covariates included physician gender, physician age, and patient gender. County, hospital, day of week, month, year, and case fixed effect were controlled for in the regression. Robust standard errors were clustered at the hospital level. PCC denotes patient-centred communication. 95% confidence intervals in brackets. ^*^ *p* < .1, ^**^ *p* < .05, ^***^ *p* < .01

### Figure S2 Propensity distribution of physician over-service in a specific hospital

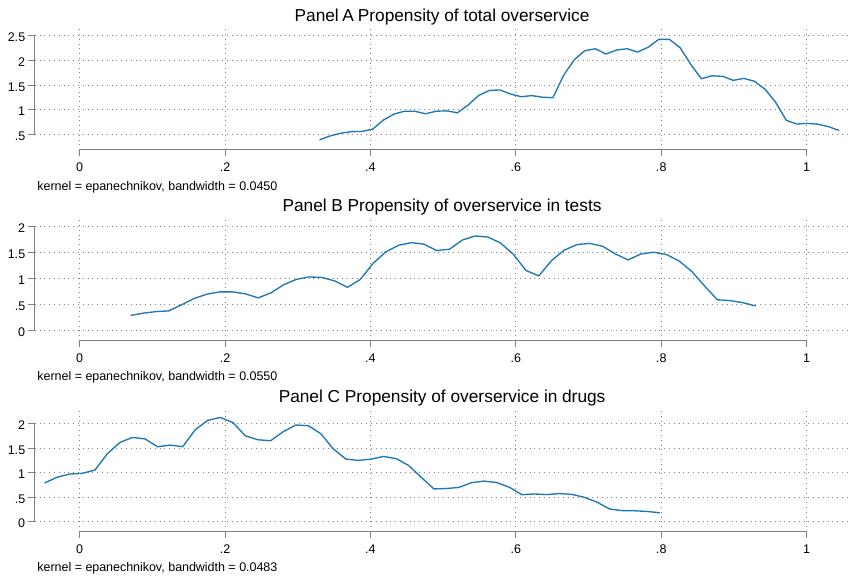

### Table S6 Comparing the quality of care across high-propensity (>=50%) and low-propensity (<50%) hospitals

|  | (1) | (2) | (3) | (4) | (5) | (6) |
| --- | --- | --- | --- | --- | --- | --- |
|  | Time | Checklist | PCC | Diagnosis | Correct drug | Referral |
| *Total over-service* | 1.838^**^ | 0.659 | 0.149 | -0.008 | 0.068 | -0.353^***^ |
|  | [0.290, 3.386] | [-0.633, 1.951] | [-3.322, 3.619] | [-0.230, 0.214] | [-0.052, 0.188] | [-0.546, -0.160] |
| *Over-service in tests* | 0.443 | -0.086 | -1.102 | -0.062 | -0.096^**^ | -0.051 |
|  | [-0.634, 1.520] | [-0.800, 0.628] | [-2.879, 0.675] | [-0.174, 0.051] | [-0.170, -0.022] | [-0.187, 0.084] |
| *Over-service in drugs* | -0.156 | 0.763^**^ | 1.957^***^ | -0.007 | 0.101^***^ | -0.129^**^ |
|  | [-1.418, 1.105] | [0.167, 1.358] | [0.583, 3.332] | [-0.156, 0.142] | [0.033, 0.170] | [-0.248, -0.009] |
| *N* | 492 | 492 | 492 | 492 | 492 | 492 |

Note: Covariates included physician sex, physician age, patient sex, hospital ownership, and participating health alliance. County, day of the week, month, year, case, and patient fixed effect were controlled for in the regression. Robust standard errors were clustered at the hospital level. PCC denotes patient-centred communication. 95% confidence intervals in brackets. ^*^ *p* < .1, ^**^ *p* < .05, ^***^ *p* < .01

### Table S7 Using different thresholds of high-propensity hospitals

40% threshold

|  | (1) | (2) | (3) | (4) | (5) | (6) |
| --- | --- | --- | --- | --- | --- | --- |
| *Total over-service* | Time | Checklist | PCC | Diagnosis | Correct drug | Referral |
| *Total over-service* | 1.977^**^ | 0.682 | 1.232 | 0.009 | 0.092 | -0.367^***^ |
|  | [0.115, 3.839] | [-0.903, 2.268] | [-2.551, 5.015] | [-0.254, 0.273] | [-0.047, 0.231] | [-0.607, -0.127] |
| *Over-service in tests* | 0.443 | -0.086 | -1.102 | -0.062 | -0.096^**^ | -0.051 |
|  | [-0.634, 1.520] | [-0.800, 0.628] | [-2.879, 0.675] | [-0.174, 0.051] | [-0.170, -0.022] | [-0.187, 0.084] |
| *Over-service in drugs* | -0.156 | 0.763^**^ | 1.957^***^ | -0.007 | 0.101^***^ | -0.129^**^ |
|  | [-1.418, 1.105] | [0.167, 1.358] | [0.583, 3.332] | [-0.156, 0.142] | [0.033, 0.170] | [-0.248, -0.009] |
| *N* | 492 | 492 | 492 | 492 | 492 | 492 |

60% threshold

|  | (1) | (2) | (3) | (4) | (5) | (6) |
| --- | --- | --- | --- | --- | --- | --- |
| *Total over-service* | Time | Checklist | PCC | Diagnosis | Correct drug | Referral |
| *Total over-service* | 0.284 | 0.620 | 0.527 | -0.016 | 0.022 | -0.209^***^ |
|  | [-0.954, 1.522] | [-0.203, 1.444] | [-1.653, 2.706] | [-0.142, 0.111] | [-0.061, 0.105] | [-0.335, -0.082] |
| *Over-service in tests* | 0.695 | 0.148 | -0.102 | -0.018 | -0.035 | -0.007 |
|  | [-0.246, 1.636] | [-0.554, 0.850] | [-1.841, 1.637] | [-0.140, 0.105] | [-0.092, 0.021] | [-0.118, 0.105] |
| *Over-service in drugs* | -0.034 | 0.774^**^ | 2.479^***^ | 0.036 | 0.117^**^ | -0.170^**^ |
|  | [-1.678, 1.610] | [0.031, 1.518] | [0.632, 4.326] | [-0.177, 0.248] | [0.018, 0.216] | [-0.324, -0.017] |
| *N* | 492 | 492 | 492 | 492 | 492 | 492 |

Note: Covariates included physician sex, physician age, patient sex, hospital ownership, and participating health alliance. County, day of the week, month, year, case, and patient fixed effect were controlled for in the regression. Robust standard errors were clustered at the hospital level. PCC denotes patient-centred communication. 95% confidence intervals in brackets. ^*^ *p* < .1, ^**^ *p* < .05, ^***^ *p* < .01

### Table S8 Instrument-variable estimates

| *Panel B* | (1) | (2) | (3) | (4) | (5) | (6) |
| --- | --- | --- | --- | --- | --- | --- |
| *IV estimates* | Time | Checklist | PCC | Diagnosis | Treatment | Referral |
| *Total over-service* | 1.737 | 1.124 | -0.437 | -0.187 | 0.074 | -0.416^***^ |
|  | [-0.697,4.172] | [-0.824,3.073] | [-5.284,4.410] | [-0.496,0.122] | [-0.085,0.232] | [-0.732,-0.100] |
| *Over-service in tests* | 1.294 | -0.194 | -2.606 | -0.078 | -0.178^**^ | -0.001 |
|  | [-0.721,3.308] | [-1.649,1.260] | [-6.234,1.021] | [-0.333,0.178] | [-0.315,-0.041] | [-0.242,0.241] |
| *Over-service in drugs* | 0.976 | 1.346^*^ | 3.721^**^ | -0.004 | 0.322^***^ | -0.427^***^ |
|  | [-1.671,3.622] | [-0.079,2.770] | [0.302,7.140] | [-0.318,0.310] | [0.209,0.434] | [-0.678,-0.176] |
| *N* | 492 | 492 | 492 | 492 | 492 | 492 |

Note: Covariates included physician sex, physician age, patient sex, hospital ownership, and participating health alliance. County, day of the week, month, year, case, and patient fixed effect were controlled for in the regression. Robust standard errors were clustered at the hospital level. PCC denotes patient-centred communication. 95% confidence intervals in brackets. ^*^ *p* < .1, ^**^ *p* < .05, ^***^ *p* < .01

### Table S9 The impact of workload on the quality of care

|  | (1) | (2) | (3) | (4) | (5) | (6) |
| --- | --- | --- | --- | --- | --- | --- |
|  | Time | Checklist | PCC | Diagnosis | Treatment | Referral |
| Busy time | -0.235 | 0.409 | 0.794 | -0.034 | 0.024 | -0.033 |
|  | [-0.990,0.521] | [-0.135,0.953] | [-0.474,2.061] | [-0.137,0.070] | [-0.039,0.088] | [-0.125,0.059] |
| *N* | 492 | 492 | 492 | 492 | 492 | 492 |
| adj. *R*^2^ | 0.22 | 0.46 | 0.20 | 0.19 | -0.01 | 0.17 |

Note: The number of waiting patients ranges from 0 to 30 with an average of 2.48. Covariates included physician sex, physician age, patient sex, hospital ownership, and participating health alliance. County, day of the week, month, year, case, and patient fixed effect were controlled for in the regression. Robust standard errors were clustered at the hospital level. PCC denotes patient-centred communication. 95% confidence intervals in brackets. ^*^ *p* < .1, ^**^ *p* < .05, ^***^ *p* < .01

### Supplement 1 Over-service in drugs and the probability of giving a correct drug

By definition, more prescribed drugs would lead to a higher cumulative probability of giving a correct drug. The process can be modelled as:

1) The prescription of different drugs is assumed to be independent.
2) On average, the probability of any prescribed drug being correct is defined as $p_{i}$.
3) The cumulative probability of giving a correct treatment for one specific case is a function of the number of prescribed drugs n.

$$P=1-\prod_{i=1}^{n} (1-p_{i})$$

The cumulative probability of prescribing at least one correct drug will converge faster when high-accuracy diagnoses are provided as $p_{i}$ increases, as visualised in *Figure 3 Panel D*.
